# Steroid Hormone Biosynthesis and Dietary Related Metabolites Associated with Excessive Daytime Sleepiness

**DOI:** 10.1101/2024.09.12.24313561

**Authors:** Tariq Faquih, Kaitlin Potts, Bing Yu, Robert Kaplan, Carmen R Isasi, Qibin Qi, Kent D. Taylor, Peter Y. Liu, Russell P. Tracy, Craig Johnson, Stephen S. Rich, Clary B. Clish, Robert E. Gerzsten, Jerome I. Rotter, Susan Redline, Tamar Sofer, Heming Wang

**Affiliations:** Division of Sleep and Circadian Disorders, Brigham and Women’s Hospital, Boston, MA, USA; Broad Institute, Cambridge, MA, USA; CardioVascular Institute (CVI), Beth Israel Deaconess Medical Center, Boston, MA, USA; Division of Sleep Medicine, Harvard University Medical School, Boston, MA, USA; Department of Epidemiology, The University of Texas Health Science Center at Houston, Houston, TX, USA; Department of Epidemiology and Population Health, Albert Einstein College of Medicine, Bronx, NY, USA; Department of Paediatrics, Institute for Translational Genomics and Population Sciences, The Lundquist Institute for Biomedical Innovation, Harbor-UCLA Medical Centre, Torrance, CA, USA; Department of Biochemistry, University of Vermont, Burlington, VT, USA; Department of Biostatistics, University of Washington, Seattle, USA; Centre for Public Health Genomics, University of Virginia, Charlottesville, VA, USA; Metabolite Profiling Platform, Broad Institute of MIT and Harvard, Cambridge, MA, USA; Department of Biostatistics, Harvard T.H. Chan School of Public Health, Boston, MA, USA

**Author notes:** Equal contribution (Tamar Sofer, Heming Wang).

**Keywords:** metabolomics, excessive daytime sleepiness, sleep, pregnenolone, cortisol

## Abstract

**Background:** Excessive daytime sleepiness (EDS) is a complex sleep problem that affects approximately 33% of the United States population. Although EDS usually occurs in conjunction with insufficient sleep, and other sleep and circadian disorders, recent studies have shown unique genetic markers and metabolic pathways underlying EDS. Here, we aimed to further elucidate the biological profile of EDS using large scale single- and pathway-level metabolomics analyses.

**Methods:** Metabolomics data were available for 877 metabolites in 6,071 individuals from the Hispanic Community Health Study/Study of Latinos (HCHS/SOL) and EDS was assessed using the Epworth Sleepiness Scale (ESS) questionnaire. We performed linear regression for each metabolite on continuous ESS, adjusting for demographic, lifestyle, and physiological confounders, and in sex specific groups. Subsequently, gaussian graphical modelling was performed coupled with pathway and enrichment analyses to generate a holistic interactive network of the metabolomic profile of EDS associations.

**Findings:** We identified seven metabolites belonging to steroids, sphingomyelin, and long chain fatty acids sub-pathways in the primary model associated with EDS, and an additional three metabolites in the male-specific analysis. The identified metabolites particularly played a role in steroid hormone biosynthesis.

**Interpretation:** Our findings indicate that an EDS metabolomic profile is characterized by endogenous and dietary metabolites within the steroid hormone biosynthesis pathway, with some pathways that differ by sex. Our findings identify potential pathways to target for addressing the causes or consequences of EDS and related sleep disorders.

**Funding:** Details regarding funding supporting this work and all studies involved are provided in the acknowledgments section.

**Research in context:** *Evidence before this study:* There is a growing recognition of the paramount importance of sleep on health and cardiometabolic disease. Excessive daytime sleepiness (EDS), one of the key common sleep treatment targets, has been linked to increased risk of mortality, hypertension, cardiovascular disease, car accidents as well as decrease in life quality, and productivity. Despite its impact on health, much remains unknown about the biological mechanisms of EDS and if those mechanisms are independent from other sleep disorders. Recent genetic evidence that shows that EDS is associated with specific genetic biomarkers supports the need to further study the underlying biology of EDS.

*Added value of this study:* Here, we used measurements of metabolites, the products and by-products of metabolism to identify the metabolomic profile of EDS. Metabolites are produced by the biological reactions within the body via proteins—themselves products of genes—and by the breakdown of external sources such as nutritional intake and breathing air pollutants. Therefore, metabolomics enables study of the effects of nutrition, environmental exposures, and genetics. In this study we aimed to identify the metabolites that were associated with excessive daytime sleepiness. Additionally, we mapped these metabolites into a publicly available online biological network of human metabolism pathways to obtain an understanding of our findings on a larger scale.

*Implications of all the available evidence:* Identifying the metabolites and pathways related to daytime sleepiness provides insights into the biological mechanisms of EDS and suggests future research opportunities to identify targets for prevention, prediction, and treatments for EDS and potentially other sleep disorders coupled with sleepiness. In this study we found 7 such metabolites—some endogenously synthesised and some obtained from dietary sources—associated with EDS. The network analysis implicated the steroid hormone biosynthesis pathway as a shared pathway underlying those metabolites, and identified linkages to key metabolites related to sleep: melatonin and cortisol metabolism.

## Introduction

Excessive daytime sleepiness (EDS) is a prevalent symptom that affects up to 33% of the population of the United States (1,2). The aetiology of EDS is heterogeneous and multi-factorial (1–5), reflecting variable contributions of insufficient sleep, some occurring secondary to sleep disorders such as sleep apnoea and circadian rhythm disorders, and others reflecting abnormalities of sleep-wake control systems such as narcolepsy and idiopathic hypersomnia (3,6). EDS frequently co-occurs with (and is often associated with increased incidence of) multiple clinical outcomes, cardiometabolic disorders such as obesity, type 2 diabetes (1), hypertension (5,7), and cardiovascular disease (2,4), impaired cognition (1,3), depression, psychiatric disorders (1–3), mortality (1–3), and reduced life quality (1–3). Notably, patients with sleep apnoea with concurrent EDS have been reported to have higher risk of cardiovascular disease outcomes than patients with sleep apnoea without EDS (4). EDS also has strong implications for public health given its contributions to work-related accidents (6), car crashes (8), and loss of productivity (2,6,9). Despite the health and social and economic impact of EDS, its aetiology and biological mechanisms are incompletely understood (2–5).

There are several challenges in evaluating the biological underpinnings of EDS. For one, although EDS is strongly linked to sleep disorders, patients may exhibit EDS even after the appropriate treatment of their respective sleep disorder (4). This indicates a potentially unique aetiology for EDS outside of those that overlap with other sleep disorders (4). Another challenge is the large inter-individual differences in EDS with overtly similar sleep disorders (3). Moreover, there are differences in prevalence of EDS across population groups and socio-economic strata that are poorly understood (9–11).

Our research approach for addressing EDS was guided by the following: First, there are genetic factors associated with EDS as shown by a recent study in the UK Biobank which identified 42 EDS related loci (3). This finding supports previous research that hypothesized that EDS is driven by biological and metabolic influences (3,6,9). Second, EDS and sleep traits are associated with cardiometabolic outcomes (4). Since cardiometabolic disorders are characterized by metabolic alterations, EDS may also be linked with metabolic alterations. The causal effects of these associations can either be directional or bi-directional and can involve shared risk factors, such as obesity (6). Accordingly, using metabolomics to quantify the association of a large number of diverse metabolites with EDS—in turn capturing the effects of genetic, metabolomic, nutritional, and environmental influences on an individual’s metabolic profile—holds considerable potential for revealing etiological insights and advancing our understanding of EDS in multi-ethic populations. Consequently, this can provide potential biological targets for intervention in order to alleviate EDS and reduce cardiometabolic risks. To our knowledge, there are a limited number of metabolomics studies on EDS and all with very small sample sizes (12).

In this study we aimed to elucidate the underlying metabolomic profile of EDS using untargeted metabolomic measurements in two large well-characterized cohorts: the Hispanic Community Health Study/Study of Latinos (HCHS/SOL) followed by replication analysis in the Multi-Ethnic Study of Atherosclerosis (MESA). In addition, we aimed to examine the links between previously identified genetic loci and metabolites implicated with EDS in order to identify the directionality of the potentially causal associations between EDS and metabolite measurements.

## Methods

### Study Designs

The HCHS/SOL is a community based prospective cohort study of 16,415 self-identified Hispanic/Latino individuals recruited from randomly selected households from geographic areas around four field centres across the United States (Chicago IL, Miami FL, Bronx NY, and San Diego CA). Baseline examination took place between 2008 to 2011. Full details regarding the sampling and study design were described previously (13,14). The baseline clinical examination provided anthropometric, biological, behavioural, and sociodemographic assessments of study participants. These included dietary intake questionnaires (two 24-hour recalls and food propensity questionnaire(15)) to derive the Alternate Healthy Eating Index 2010 (AHEI-2010)(16), physical activity assessment based on a modified version of the Global Physical Activity Questionnaire to derive total physical activity in a week (MET-min/day)(17), the Center of Epidemiologic Studies Depression Scale (18) to assess depression (CESD10), the State-Trait Anxiety Inventory (STAI10) (19) to assess anxiety, self-report medication usage, smoking (never, former or current), alcohol use (never, former or current), hypertension status (based on medication use, self-report, and if systolic or diastolic blood pressure was greater than or equal to 140/90), and diabetes status as defined by the American Diabetes Association (20). In addition, several sleep traits were collected using self-report sleep questionnaires (Women’s Health Initiative Insomnia Rating Scale (WHIIRS)(21), and the Epworth Sleepiness Scale (ESS)(22). Participants also reported bedtime and wake time during weekday and weekend days, from which weighted average sleep duration was computed ((5×average weekday sleep duration + 2×average weekend sleep duration) / 7). Sleep apnoea severity was assessed using respiratory event index (REI, 3% desaturation) scored from in-home sleep apnoea test using the Apnea Risk Evaluation System (ARES Unicorder, Advanced Brain Imaging, Carlsbad CA) (23). 7-day actigraphy derived sleep metrices were collected and scored in the Sueño ancillary study (N= 2,252) between 2010 and 2013 using Actiwatch Spectrum (Philips Respironics) (24). Using this data, sleep efficiency and sleep mid-point were evaluated in this subsample.

Excessive daytime sleepiness was defined based on the ESS questionnaire (22). Briefly, ESS includes 8 items regarding the likelihood of dozing off in different scenarios during the day on a scale from 0 to 3. An overall ESS was derived from the individual item score and ranges from 0 to 24, with higher scores corresponding to higher sleepiness, wherein a score of 11 and above is commonly use to define EDS (6,22). The ESS was available in 13,820 individuals in the HCHS/SOL study. Details regarding the MESA study design and measurements, including metabolomics methods, are provided in the supplementary materials.

### Metabolomic Measurements

In HCHS/SOL, serum blood samples were collected at baseline (between June 2008–July 2011) for all 16,415 individuals. Of these samples, 6,372 were sent to Metabolon for the quantification of metabolomic measurements. Metabolomic quantification was performed in two batches, in 2017 and 2021 respectively, by Metabolon Inc. (Durham, NC) using their Discovery HD4 Ultra-High-Performance Liquid Chromatography tandem mass spectrometry platform (25). Samples were stored in −70°C freezers after collection until assayed (26). Overall, 4004 samples were measured in the first batch and 2,368 were measured in the second batch. The samples of 152 individuals were used for quantification in both batches. For these repeated measurements we only included the measurements in batch 2. In addition, we excluded samples of two individuals that were quantified twice within the first batch and 37 individual samples quantified twice within the second batch. After these exclusions, 3,850 samples were available in the first batch and 2,330 samples were available from the second batch. Therefore, after including only one sample per individual, the total number of samples included was 6,071. After the metabolomics quantification was performed, a total of 879 metabolites were quantified in the first batch and 1264 metabolites were measured in the second batch. However, two metabolites were excluded in the first batch as they were not detected in the included individuals with ESS data. Finally, we selected the overlapping metabolites between the two batches—877 metabolites—for statistical analysis.

### Metabolomics Data Processing in the HCHS/SOL study

To mitigate the effect of outlier metabolite measurements we winsorized metabolite levels that were 5 standard deviations from the mean.

Multiple imputation using chained equations was used to impute missing values in all metabolites to produce 5 imputed datasets for batch 1 and batch 2 separately. The methodology applied was described in detail in previously published work (27). The two batches were then merged into 5 imputed datasets. Finally, all metabolites were log transformed and centred and scaled to have a mean of 0 and a standard deviation of 1.

### Multivariable linear regression analysis

We conducted a series of sequential covariate-adjusted models to assess the metabolite associations with EDS. Summary of all model designs and a directed acyclic graph are provided in Supplementary Table 1 and Supplementary Figure 1. Multivariable linear regression or logistic regression was performed with continuous and dichotomized ESS (above vs below a score of 10) for each metabolite separately. For the minimally adjusted model (Model 1) we adjusted for age, BMI, gender, and 7-level classification of self-reported Hispanic background (Dominican, Central American, Cuban, Mexican, Puerto Rican, South American, or “More than one/Other” heritage). Sample weighting of the study design was accounted for as described (13). For the primary model (Model 2) we additionally adjusted for self-reported alcohol, cigarette use and weekly physical activity. A third model (Sleep Traits Model) was conducted to examine if the associations were specific for EDS or reflected other measured sleep traits. This model adjusted for short and long sleep duration (<7 and >=9 hours) (28), insomnia (continuous), and sleep apnoea (continuous), in conjunction and individually in addition to the previously listed primary confounders. All analyses were repeated stratified by sex. To adjust for multiple testing, we used the method described by Li and Ji et al (29) to calculate the independent number of metabolites. Accordingly, there we calculated 390 independent metabolites and set the significance cutoff to 0.05/390 = 0.00013.

Several additional sensitivity analyses were performed. First, we accounted for the effect of dietary effects by adjusting for the AHEI-2010 score in the diet model. Second, we accounted for potential confounding due to hypertension, statin usage, and diabetes in the comorbidities model. We performed sensitivity analyses independently adjusting for anxiety (STAI10) and depression (CESD10), as well time of blood draw in addition to the minimally adjusted model confounders. An additional analysis was performed adjusting for the available sleep medications usage and shift work status data in the Sueño ancillary.

In line with the prior EDS GWAS (3), a crude analysis was performed to examine the association between the statistically significant metabolites with actigraphy derived sleep data in the Sueño ancillary subset to understand their effects on subtypes of EDS (sleep insufficiency vs sleep propensity). Specifically, metabolites linear associations with mean bedtime and wakeup times in the weekday and weekend, mean daily sleep duration, and average daily sleep efficiency (proportion of sleep duration in the total rest period) in individuals with at least 4 validated actigraphy days (N= 1,852) were analysed without adjustment for other variables

### Network Analysis and Pathway analyses

To provide context to the relationship between the metabolite associations with EDS across biological pathways, we used a three-way approach to produce an interactive network. This included (i) a method to capture statistically significant correlations between the measured metabolites, , (ii) a pathway enrichment analysis and (iii) explication of a biologically driven network between the metabolites and known reactions and pathways. First, gaussian graphical modelling (GGM) using sparse inverse covariance estimation with the graphical lasso (30,31) was used to calculate the correlations between the metabolites measured in the HCHS/SOL study. To reduce the number of irrelevant metabolites in relation to EDS, we selected metabolites that were associated with the outcome in each of the linear regression models (from either Model 1-2 for either pooled and sex stratified analyses) and had a P value < 0.005. This step helps reduce the number of irrelevant metabolites to the outcome and in turn unclutters the network and reduces overfitting of metabolite connections. Results of the GGM analysis were then imported into Cytoscape(32) to visualize the metabolites correlations. Second, the selected metabolites and a selection of genes previously found to be associated with EDS (3) were supplied to the MetaboAnalyst 6.0 online metabolomics analysis suite (33,34) to perform a pathway analysis based on global testing (35) and “Metabolite Set Enrichment Analysis”. Specifically, we performed pathway analysis using the metabolites only, followed by a joint pathway analysis using both EDS associated genes (3) and metabolites. Pathway enrichment analysis was performed to identify the most enriched pathways. Results of the joint pathway analysis and the pathway enrichment analysis were then combined with the GMM network developed in the first step. Third, additional annotations were added to the network within Cytoscape using the Metscape plug-in (36). Metscape provided annotations for the metabolites and added nodes for relevant biological reactions and genes involving pathways of the metabolites based on the data from the Kyoto Encyclopaedia of Genes and Genomes (KEGG) and The Edinburgh Human Metabolic Network (37) to our network. Moreover, biologically relevant intermediate metabolites involved in these reactions and pathways, but not measured in our dataset, were added to the network and given a unique labelling to distinguish them from the measured metabolites.

These steps resulted in a fully interactive webpage (Cytoscape JS) of networks that combined the GGM approach as well as biologically driven pathway and enrichment analyses.

### Mendelian randomization analysis

Mendelian randomization (MR) analysis was performed using the TwoSampleMR R package (Version 0.6.0) (38). We used the metabolite quantitative trait locus (metabQTL) data from the Trans-Omics in Precision Medicine (TOPMed) program Phase 1 TOPMed metabQTL produced as part of TOPMed Metabolomics standard operating procedure work. This work included 16,359 individuals across 8 TOPMed studies using the Broad Institute and Beth Israel Metabolomics Platform or Metabolon platform. Full details are provided in the Supplementary materials. Summary statistics for EDS were obtained from the GWAS of the UK Biobank (N= 452,071) (3).

MR analysis was performed using the MR Egger, weighted median, inverse variance weighted, simple mode, and weighted mode methods using statistically significant metabolites in all our analyses that were also available in the TOPMed metabQTL as the exposure and EDS as the outcome. All analyses used default minor allele frequency cutoff (MAF > 1%). Heterogenicity testing was performed using the Cochran’s Q statistic and directionality testing was performed using the MR Steiger directionality test (39).

### Ethics

This work was approved by the Institutional Review Board of Brigham and Women’s Hospital. HCHS/SOL assessments were approved by the Institutional Review Boards of the participating institutions. This study was approved by the institutional review boards of all MESA field centres. All participants provided written informed consent in both studies.

## Results

### Population characteristics

After exclusions of duplicate metabolomic measurements and individuals with missing ESS or BMI, our study included 6,071 individuals of Hispanic/Latino background (Figure 1). The characteristics of the study are summarized in Table 1. The distribution of individuals from each field centre was similar and the majority of individuals self-identified as Mexicans (32.5%). The mean age was 48.28 years and consisted of 60% women (N=3,635). Average BMI was approximately 30 in both sexes. Overall mean of ESS was 5.79 and was slightly higher score in men (5.98). Using the 10-point cutoff, 957 (15%) individuals were classified as having EDS (of which 58% were women). Regarding other assessed sleep traits, 11% had moderate sleep apnoea (Respiratory Event Index>15 events/hr; 59% of which were men), mean sleep duration was 7.91 hours, and the average insomnia score (WHIIRS) was 7.46. Finally, the reported average time of blood sample collection was around 10am.

**Figure 1:**
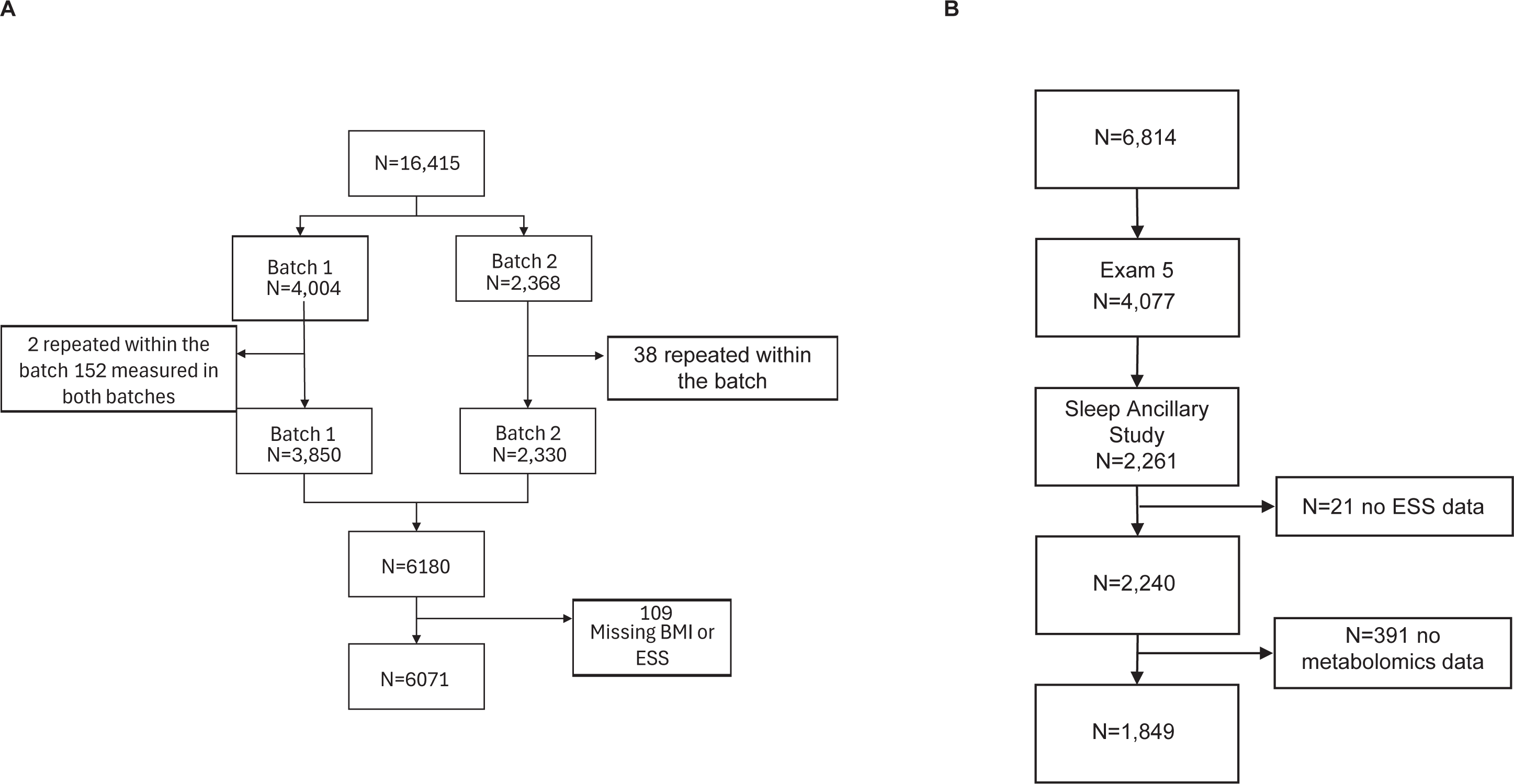
Inclusion criteria for the current study from the HCHS/SOL study

In total 877 metabolites were available for all individuals in this study. These metabolites included 320 lipids (36.4%), 175 amino acids (19.9%), 139 unannotated metabolites (15.8%), and 120 xenobiotics (13.6%)—which includes metabolites derived from medications, environmental exposures, and nonessential dietary sources such as caffeine. The remaining metabolites belonged to peptides, nucleotides, vitamins, carbohydrates, and partially characterized molecules.

### Primary metabolite association analysis

The minimally adjusted model (Model 1), adjusting for age, sex, BMI, and Hispanic background, identified 7 metabolites (P<0.00013) negatively associated with the ESS on the continuous scale (Table 2; Supplementary Table S2, Supplementary Figure 2). Of these metabolites, 5 belonged to lipids, specifically, long chain polyunsaturated fatty acids (PUFA), steroids (Pregnenolone and Corticosteroids), and sphingomyelin. The remaining 2 metabolites (X-11470 and X-11444) were unannotated, and their pathway was not available in the Metabolon database. All effect directions were negative (i.e. higher ESS was associated with lower metabolite levels) and ranged between - 0.34 and −0.4. After further adjustment for smoking, alcohol use, and physical activity (MET-min/day) in the primary model (Table 2; Supplementary Table S2), the effect estimates and their direction remained consistent and only docosadienoate was no longer statistically significant (p = 1.41×10^-04^). The diet model, adjusting for AHEI-2010 score (Table 2; Supplementary Table S2), had minimal changes to the effect sizes and directions as well. However, the associations of pregnenediol sulfate, tetrahydrocortisol glucuronide, and X-11444 were not statistically significant in this model. Finally, after adjusting for comorbidities (Table 2; Supplementary Table S2) the effect estimates remained consistent with the previous models, however, pregnenediol sulfate and X-11444 were no longer significant. In addition to the associated metabolites, several sphingomyelins and steroid related metabolites were borderline statistically significant (P value ∼ 0.0001) in all models (Supplementary Figure 3 and 4).

### Sleep traits analysis and sensitivity analyses

After adjusting for sleep traits (insomnia, OSA, and sleep duration) in addition to the covariables used in the primary model, diet, and comorbidities models respectively four of the metabolites reported in the primary model remained statistically significant with slightly stronger effect sizes in the same direction (negative direction) (Table 3 and Supplementary Tables S3, S6, and S7). Effect sizes and directions remained consistent after adjusting for diet and comorbidities. Individual adjustment of sleep traits (insomnia, OSA, long sleep, and short sleep) did not alter the results. The only exception was the association of adrenate (22:4n6) with EDS which was only significant after adjusting only for insomnia along with comorbidity confounders. For two sensitivity analyses, neither the first sensitivity analysis (adjusting for the minimally adjusted model in addition to blood draw timing, depression and stress scores) nor the second sensitivity analysis (adjusting for shift work and sleep medications in the Sueño ancillary) altered the results for the primary analyses (Supplementary Table S9 and S10).

### Sex stratified analyses

The metabolomic profiles in the sex-stratified subpopulations exhibited distinct patterns in comparison to the primary analyses. First, no significant associations were found in any female-specific analyses (Supplementary Table S5). Second, the analysis for males identified 6 metabolites associated with EDS in the minimally adjusted analysis (Table 4; Supplementary Table S4). Three of these metabolites were previously found to be associated with EDS in the pooled analyses with both men and women: docosadienoate (22:2n6), X-11470, and tetrahydrocortisol glucuronide. The other 3 metabolites were associated with EDS only in males: two glycerophospholipid (GPC) phosphatidylcholines (1-stearoyl-2-arachidonoyl-GPC (18:0/20:4) and 1-palmitoyl-2-arachidonoyl-GPC (16:0/20:4n6)) and tyramine O-sulfate. After adjusting for variables in the primary model (model 2), X-11470 and tetrahydrocortisol glucuronide were no longer significant. Adjusting for diet additionally excluded tyramine O-sulfate from the significance threshold. Finally, compared to the original 6 metabolites identified in the minimally adjusted analysis, in the comorbidity adjusted analysis tetrahydrocortisol glucuronide and X-11470 were no longer significant and the remining associations maintained the same effect direction and similar effect sizes. However, an additional metabolite, phenylacetylcarnitine, was significantly associated with EDS in males in the comorbidity adjusted analysis only.

### Association with Actigraphy data

Crude analysis was performed between the ten significant metabolites associated with EDS identified in the prior analyses and the actigraphy measures in the Sueño subsample (summarized in Table 5). Notably, dihomo-linoleate (20:2n6) and sphingomyelin (d18:2/16:0, d18:1/16:1), which were associated with lower EDS, were associated with both longer sleep duration and higher sleep efficiency, a pattern which reflects a higher sleep propensity subtype. Pregnenediol sulfate was associated with shorter sleep duration and lower EDS. Finally, tetrahydrocortisol glucuronide was associated with lower sleep efficiency and reduced EDS, which reflects a mixed association of reduced sleepiness during daytime but reduced sleep efficiency during nighttime.

### Network and pathway enrichment

Based on the correlations and metabolic network for the metabolites associated with EDS, we found biological links with several pathways. Primarily, steroid hormone biosynthesis was at the centre of biological pathways associated with EDS (Figure 2, Online supplementary materials, https://tofaquih.github.io/Metabolomics-Excessive-Daytime-Sleepiness/). This was confirmed by the pathway enrichment analysis. The enrichment was driven by pregnenediol sulfate, tetrahydrocortisol glucuronide, and dihomo-linoleate (20:2n6) as well as the *CYP1A1*, *CYP1A2*, and *CYP7B1* genes identified from the previous GWAS on EDS (3). These metabolites were also linked to cortisol and cortisone metabolism. In turn, dihomo-linoleate (20:2n6) was correlated with 11 beta-hydroxyandrosterone glucuronide (i.e. 6-hydroxymelatonin), linked to the metabolism of melatonin and to the aforementioned genes. Tyramine O-sulfate also shared a biological reaction and enzyme related to the CYP genes and melatonin pathways. Finally, sphingomyelin (d18:2/16:0, d18:1/16:1) was as expected, correlated with other species of sphingomyelin. Those sphingomyelins seem to be indirectly correlated with pregnenolone steroids as well.

**Figure 2:**
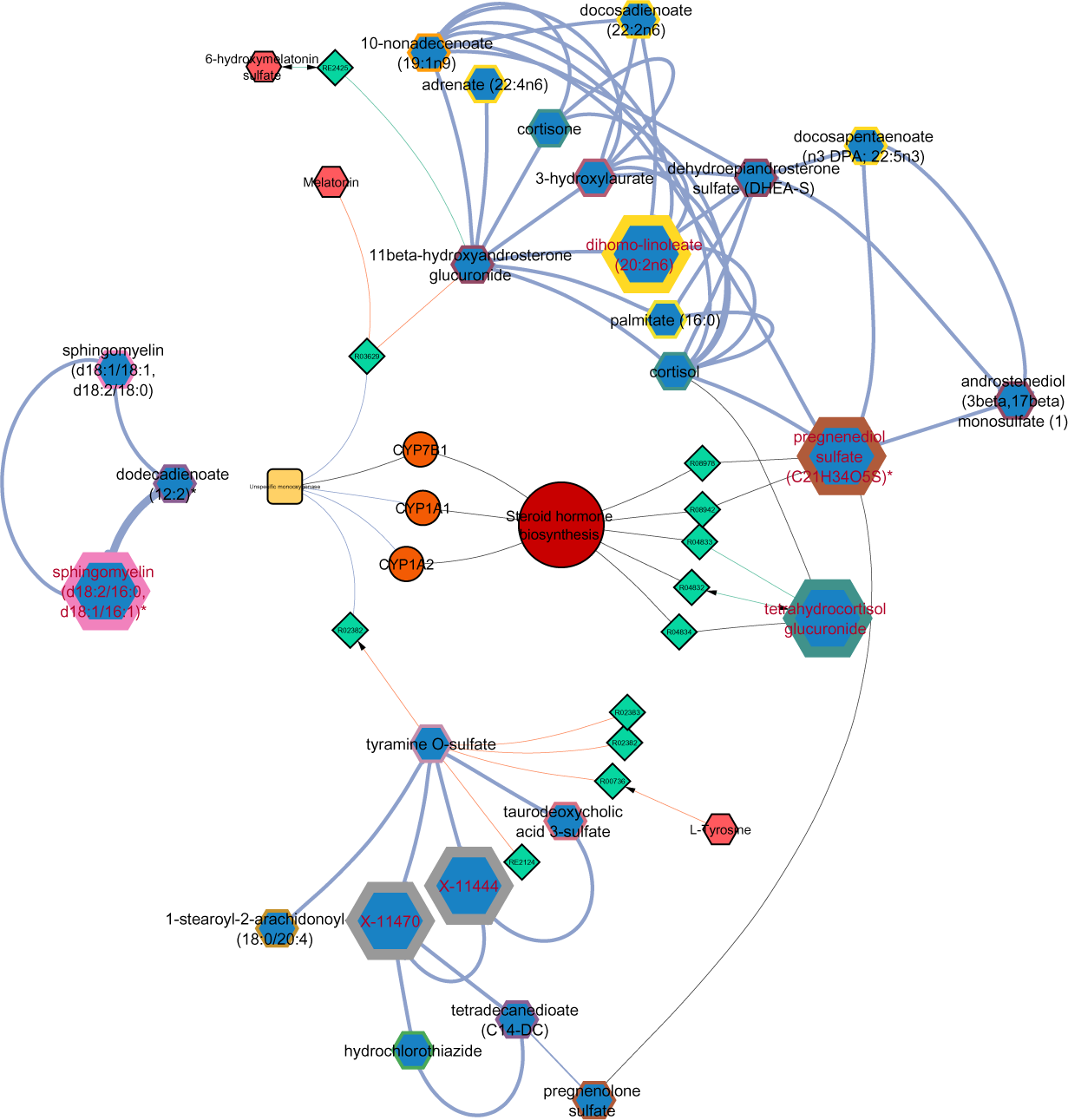
Network of Metabolites and related pathways associated with EDS.

### Replications in MESA

Of the 11 statistically significant metabolites observed in HCHS using the Metabolon platform, only four were available in the MESA samples assayed with the Broad platform: pregnenediol sulfate (C21H34O5S), pregnenolone sulfate, dihomo-linoleate (20:2n6), 1-stearoyl-2-arachidonoyl-GPC (18:0/20:4), or sphingomyelin (d18:2/16:0, d18:1/16:1). None of these reached statistical significance (P<0.05), although the direction of the associations was consistent with the results from the HCHS/SOL study apart from dihomo-linoleate (Supplementary Table S8).

### Mendelian randomization Associations

MR analysis was performed for eight metabolites that were reported in the analyses models, had a an identifiable chemical name, and had available genetic summary statistics required for the MR analysis: tetrahydrocortisol glucuronide, pregnenetriol sulfate, docosadienoate (22:2n6), dihomo-linoleate (20:2n6), tyramine-O-sulfate, 1-stearoyl-2-arachidonoyl-GPC (18:0/20:4), 1-palmitoyl-2-arachidonoyl-GPC (16:0/20:4n6), and phenylacetylcarnitine. We identified statistically significant associations (p<0.05) for pregnenediol sulfate (inverse variance weighted (IVW) beta= −0.017, p= 0.044), tetrahydrocortisol glucuronide (IVW beta= −0.013, p= 0.047), and tyramine-O-sulfate (IVW beta= 0.03, p= 0.004), consistent with a potential causal (protective) association between these metabolites and EDS. Full results are provided in Table 6. No heterogeneity or horizontal pleiotropy were detected, and effect direction was confirmed for these results by the Steiger directionality test.

## Discussion

In this study we identified seven metabolites associated with EDS, and an additional four associated specifically in the male stratified analysis. These metabolites were primarily driven by steroid hormonal biosynthesis pathways as indicated in our pathway enrichment analysis and network. First, we report two specific pregnenolone steroids(40), pregnenediol sulfate and tetrahydrocortisol glucuronide, with higher levels associated with less sleepiness. These associations were consistent with a causal association in our mendelian randomization analysis. and maintained the same effect direction as our linear regression analyses. These metabolites are involved in the pathways required for conversion to glucocorticoids, neurosteroids, androgens, and cortisol (41,42). Furthermore, pregnendiol sulfate appears to have an immunosuppressive role in certain infections(43) and tetrahydrocortisol glucuronide is a major metabolite of cortisol and a marker of adrenocortical activity. Pregnendiol sulfate is also acutely regulated by the pituitary hormone ACTH, which strongly implicates its regulation by the adrenal axis (40). Together, these data suggest that the protective association between these adrenal steroids and EDS reflects an immunosuppressive role of adrenocortical activation – a role that is consistent with known anti-inflammatory effects of glucocorticoids. This interpretation is consistent with data that we and others have generated showing that greater EDS in OSA is related to inflammation (44,45). In addition, cortisol-testosterone balance is important for sleep and general health. Insufficient sleep duration has been reported to disrupt the hormonal balance, leading to decreased testosterone and increased cortisol levels (46). The role of the adrenal gland with EDS is further supported by reported associations between androstenedione, which partakes in circadian regulation in the adrenal gland, with ESS in men (47).

Notwithstanding the predominately adrenal origin of pregnenediol sulfate and tetrahydrocortisol glucuronide, such steroids may not be of *exclusively* adrenal origin, so a role of ovarian activation should also be considered particularly because steroids of gonadal origin exhibit differential circadian and sleep regulation in a sex-specific manner (45,48). Notably, clinical trials (mostly with male subjects) and observational studies have reported progesterone steroids—produced from pregnenolones—and their derivatives partake in inducing sleep, act as a sedative, increase somnolence, reduce sleep latency, reduce sleep apnoea and sleep breathing disorders, and increase non rapid eye movement sleep (41,43). In women, progesterone concentrations are reduced post menopause (41,42). This reduction, which occurs after menopause, is associated with longer sleep latency and more difficulty maintaining sleep compared to premenopausal women (49–51); while administration of progesterone in postmenopausal (52) women showed reduced wakefulness during sleep based on a EEG clinical trial (53). In addition, regulation of progesterone levels (and sex steroid hormones) has been linked to circadian rhythm (48) and menstrual phases in premenopausal women (45). Although, progesterone levels have been hypothesized to be protective of sleep apnoea and sleep disordered breathing in premenopausal women (54,55), the prevalence of EDS has been reported in some studies to be higher in women than in men (56–58), while other studies reported that the EDS was specifically higher in post/peri-menopausal women.

Another property of progesterone is its involvement with γ-Aminobutyric acid (GABA) neurotransmitter. Progesterones (such as pregnenediol sulfate) and glucocorticoids (such as tetrahydrocortisol) have been reported to exert an agnostic effect on the GABA_A_ receptor (59–61). GABA_A_ receptors are major targets of sleep medications such as benzodiazepine used for promoting sleep in patients with insomnia, anxiety, and other neurological disorders (62,63). Indeed, the effect of progesterone on sleep quality in both men and women has been suggested to be due to an interaction between progesterone and the GABA_A_ (similar to the action of benzodiazepines) (41,53,59,64). Interestingly, after adjustment for diet quality, both metabolites were no longer significantly associated with EDS. This is in line with the influence of diet and cholesterol in the biosynthesis of these metabolites (65). Actigraphy analysis for pregnenediol sulfate actigraphy metrices further indicates an association with of reduced sleepiness and shorter sleep duration, indicating a sleepiness resilience despite shorter sleep duration.

We did not detect significant associations between progesterone steroids or any other metabolites in analyses restricted to females. However, the effect direction was consistent with the primary results. Thus, our findings from the linear regression and MR analyses for pregnenediol sulfate and tetrahydrocortisol glucuronide are indicative of a larger role of glucocorticoid and adrenal gland related hormones with EDS in men. Lack of significant findings for women, despite progesterone steroids role as sex hormones particularly in women (41), could be due to the complex regulation and menstrual cycle variation of progesterone levels in women and the inclusion of women with different menopausal stages, which may have obscured potential associations when examining the data stratified by sex. Future studies designed specifically to examine women’s health are needed to further understand sex-specific risk factors for EDS.

Second, we report two long chained fatty acids obtained primarily from dietary sources: omega-6 fatty acid dihomo-linoleate (20:2n6) (also known as dihomolinoleate or DGLA) and docosadienoate (22:2n6) (i.e., docosadienoate). Accordingly, these metabolites are involved in fatty acids metabolism and are upstream of docosahexaenoic acid (DHA) and other known fatty acids (66). Interestingly docosadienoate was found to be associated with EDS when we adjusted for diet and comorbidities. Although we did not find studies that found an association between these two metabolites with EDS specifically, previous sleep research has examined the associations of fatty acids intake in general with sleep quality and sleep disorders. Long chain fatty acids are known to be involved in the production of melatonin and DHA supplementation has been suggested to improve sleep quality (67,68). For example, a randomized control trial in children between the ages of 7-9 (N=395) which concluded that higher levels of docosahexaenoic acid— where associated with improved sleep in the children (69). Omega-6 fatty acids have also been shown to be associated with sleep quality and slow wave sleep in obese individuals (66) and consumption of omega-3 and omega-6 fatty acids rich foods was also associated with improved sleep efficiency(70). Regarding the specific fatty acids reported in our study, dihomolinoleate was associated with sleep midpoint (71) and both dihomolinoleate and docosadienoate were linked to, and possibly regulated by, circadian rhythm (72)<u>. The sources of these metabolites (such as vegetable oils, nuts, seeds and fatty fish</u>(73)<u>) are abundant in Mediterranean-type diet, which in turn has been associated with several positive sleep outcomes, including higher sleep quality</u>(74–76) <u>and reduced daytime sleepiness</u>(77). Overall, our findings align with the literature regarding the benefits of long chain fatty acids to improving sleep and, in this particular study, by reducing sleepiness. Furthermore, our findings corroborate the role of long chain fatty acids for the biosynthesis of melatonin, as indicated by their connection with pregnenolone steroids and melatonin in the pathway enrichment network and analysis. Finally, actigraphy analysis implicates there two metabolites with a reduced sleepiness and longer sleep duration.

Third, we have identified sphingomyelin (d18:2/16:0, d18:1/16:1) to be associated with reduced EDS as well as four sphingomyelins—highly correlated with (d18:2/16:0, d18:1/16:1)— that were borderline significant. Sphingomyelin and sphingolipids are integral in the biosynthesis and regulation of steroid hormones and cortisol (78), particularly in adrenal cortex (78,79). In addition, sphingolipid biosynthetic regulates the steroidogenic gene expression and activity (79). Thus, our findings and pathway analysis is in line with biological pathway of sphingomyelin species in relation to pregnenolone steroids and cortisol and their subsequent association with EDS. Sphingomyelin has also been reported in previous metabolomic study on sleep disordered breathing, hypertension, and type 2 diabetes (80).

Regarding the sex stratified analysis, an interesting association was that of tyramine O-sulfate with EDS in males. In addition, mendelian randomization analysis for this metabolite further suggests a potential causal association with EDS. Tyramine O-sulfate is a secondary metabolite of tyramine, a monoamine that can occur endogenously or can be obtained from dietary sources (81,82) such as fermented foods, wine, beer, coffee, cheese, among others (82). Tyramine levels are particularly higher in overripe and spoiling food, thus high levels of tyramine are an indication of poor food quality or poor food storage (82). As a monoamine, tyramine is related to, and partakes in the biosynthesis of, monoamine neurotransmitters including serotonin, dopamine, histamine, noradrenaline, adrenaline among others (81,83). Numerous research studies have reported the role of monoamines on sleep. For instance, levels of these monoamine have been linked with sleep-wake stages and circadian rhythms (83–86). Specifically, monoamines levels were higher during sleep deprivation and remained at high levels during sleep recovery (86). In addition, monoamines are generally lower during wake stages compared to sleep stages (86), and therefore consumption of tyramine rich foods could lead to higher levels of monoamines, such as adrenaline can in turn can negatively affect sleep (87). In relation to melatonin, some tyramine reactions involve the monoamines oxidase enzymes (such as monoamine oxidase A and B), which in turn are related to flavoprotein. Our network analysis also indicates that tyramine reactions shared the enzymatic group (unspecific monooxygenase/flavoprotein monooxygenase) with melatonin, specifically 6-hydroxymelatonin. Interestingly a study on mice examining the inhibitory and regulatory effects of tyramine on monoamine oxidase reported an accumulation of 6-hydroxymelatonin, along with tyramine, histamine, and taurine, in trace amine-associated receptor 1 (TAAR1) knock out mice (88). As noted by that study, melatonin, histamine, and taurine are associated with sleep regulation, circadian rhythms (88,89). Additionally, *TAAR1* has been associated with the regulation and increase wakefulness in animal models (88,89) and a potential drug target for sleep disorders (90). Another shared aspect between melatonin and tyramine are the CYP genes: *CYP1A1*, *CYP1A2*, and *CYP7B1*. These three genes were included in our network analysis as they were previously associated with EDS (3). These genes are also implicated with unspecific monooxygenase/flavoprotein monooxygenase enzymes and metabolism, which involve monoamines such as tyramine.

This body of evidence as well as our findings suggests a role of tyramine in sleepiness and possibly sleep regulation. However, the exact biological pathway—whether via monoamine oxidase or *TAAR1* inhibition, or production of neurotransmitter monoamines, or by affecting melatonin levels—remains unclear and beyond the scope of our study. Further studies to elucidate this biological mechanism are required.

The two unannotated metabolites associated with EDS in the primary and stratified models, X-11470 and X-11444, were directly found to be correlated with tyramine O-sulfate in our network analysis. Based on a recent metabolome genome wide association study, both metabolites shared the same locus and “metabotype” (i.e. a cluster of metabolites sharing at least one genetic signal) on the *SRD5A2* gene along with 10 androgenic steroids related metabolites (91). This was further supported by the direct feedback from the Metabolon structure analysis that predicted, based on their chemical properties, that they were likely steroid glucuronides. Previous GWA studies reported associations between *SRD5A2* locus has been associated with testosterone levels (92) and hair loss/balding (92,93) in men. These findings and correlation of X-11470 and X-11444 with tyramine O-sulfate could indicate the possible connection between androgenic steroids in men with tyramine and sleepiness.

Finally, two GPCs (1-stearoyl-2-arachidonoyl-GPC (18:0/20:4) and 1-palmitoyl-2-arachidonoyl-GPC (16:0/20:4n6)) were associated with EDS in the men stratified analysis. GPCs are involved in a wide range of biological functions, including biological involvement in pathways encompassing dihomo-linoleate and docosadienoate (94,95). Similarly, they are derived from dietary sources, such as fish, milk fat, and eggs, that have been associated with improved sleep quality(70,96,97). Thus, their association with EDS is likely related to the fatty acids associations we have identified and share similar dietary sources. In addition, GPCs specifically were reported in previous sleep studies. GPCs found to be associated with blood pressure in obstructive sleep apnoea in patients (94) and were increased in during sleep deprivation (98). 1-stearoyl-2-arachidonoyl-GPC specifically was associated with sleep disordered breathing as well (80).

Our study had several strengths. We employed metabolomics analysis, examining a broad range of metabolites across various biological pathways. This comprehensive approach, combined with genomics and pathway analysis, yielded novel insights into EDS, particularly for the Hispanic/Latino population. In addition, the scale and sample size of this study, is to our knowledge, the largest metabolomic study on EDS to date (12). However, we were not able to replicate the small set of available metabolites in the MESA study. Lack of replication in the MESA study can be attributed to several factors. First, the sample available for the replication was small which reduced the power of the analysis. Second, detected metabolites were between the studies were inconsistent due to the different metabolomic measurement methodologies and platforms used. For example, 1-stearoyl-2-arachidonoyl-GPC measurement was only available in 1,327 individuals while dihomo-linoleate was available in 1839 individuals. Third, the difference in population characteristics, such as age—mean age of 48 in HCHS/SOL compared to 68 years old in MESA) and ethnicity, can affect EDS and sleeping habits in general, as well as metabolomic profiles (99). Finally, although the measurements in HCHS/SOL and MESA were performed using platforms utilizing chromatography and mass spectrometry techniques, the process, overall methodology, and final quantification of the metabolites are not identical. This is an important factor that reduced our replication power as well.

## Conclusions

In conclusion, we have identified steroid hormone biosynthesis as the primary driver for the metabolite associations with EDS via pregnenediol sulfate and tetrahydrocortisol metabolites and potentially the GABA_A_ receptor. In addition, our findings suggest the role of dietary derived metabolites, specifically fatty acids, sphingomyelin, GPC, and tyramine, on sleepiness. Tyramine in particular could be of interest in future studies due to it is association with the TAAR1 receptor, a potential drug target for sleep disorders. These findings were supported by the previous GWAS on EDS and pathway and enrichment analysis. Overall, these metabolites and pathway sheds light on the metabolomic profile of EDS in the Hispanic/ Latino population, EDS profile differences in males, and EDS in metabolomic profile in general.

## Data Availability

HCHS/SOL study and MESA pseudonymized data are available via controlled-access application to dbGaP (study accession phs000810 and phs003288.v1.p1) or via approved data use agreement with the Data Coordinating Centre of the HCHS/SOL (University of North Carolina) and MESA (University of Washington). For more details see https://sites.cscc.unc.edu/hchs and https://internal.mesa-nhlbi.org/. Individual WGS data for TOPMed and metabolomic data for MESA can be obtained by application to dbGaP with accession number phs001416.v2.p1.

https://internal.mesa-nhlbi.org/

https://www.ncbi.nlm.nih.gov/gap/

## Contributors

Tariq Faquih.: conceptualization, formal analysis, methodology, visualization, writing – original draft.

K. P.: Writing – review & editing

Bing Yu: Writing – review & editing

Robert Kaplan: Project administration, Writing – review & editing

Carmen R Isasi: Project administration, Writing – review & editing

Qibin Qi: Writing – review & editing

Susan Redline: Supervision, Writing – review & editing

Tamar Sofer*: Funding acquisition, Supervision, Conceptualization, Writing – review & editing

Heming Wang*: Funding acquisition, Supervision, Conceptualization, Writing – review & editing

## Acknowledgments

This work was supported by the National Institute of Health (NIH) grants R01HL153814 (to H.W.), R01HL161012 (to T.S.). Support for metabolomics data was graciously provided by the JLH Foundation (Houston, Texas). The Hispanic Community Health Study/Study of Latinos was carried out as a collaborative study supported by contracts from the National Heart, Lung, and Blood Institute (NHLBI) to the University of North Carolina (N01-HC65233), University of Miami (N01-HC65234), Albert Einstein College of Medicine (N01-HC65235), Northwestern University (N01-HC65236), and San Diego State University (N01-HC65237). The following Institutes/Centers/Offices contribute to the HCHS/SOL through a transfer of funds to the NHLBI: National Center on Minority Health and Health Disparities, the National Institute of Deafness and Other Communications Disorders, the National Institute of Dental and Craniofacial Research, the National Institute of Diabetes and Digestive and Kidney Diseases, the National Institute of Neurological Disorders and Stroke, and the Office of Dietary Supplements. The authors thank the staff and participants of HCHS/SOL for their contributions.

The MESA projects are conducted and supported by the National Heart, Lung, and Blood Institute in collaboration with MESA investigators. Support for MESA is provided by contracts 75N92020D00001, HHSN268201500003I, N01-HC-95159, 75N92020D00005, N01-HC-95160, 75N92020D00002, N01-HC-95161, 75N92020D00003, N01-HC-95162, 75N92020D00006, N01-HC-95163, 75N92020D00004, N01-HC-95164, 75N92020D00007, N01-HC-95165, N01-HC-95166, N01-HC-95167, N01-HC-95168, N01-HC-95169, UL1-TR-000040, UL1-TR-001079, and UL1-TR-001420, UL1TR001881, DK063491, and R01HL105756. A full list of participating MESA investigators and institutes can be found at http://www.mesa-nhlbi.org. The authors thank the other investigators, the staff, and the participants of the MESA study for their contributions. Molecular data for the Trans-Omics in Precision Medicine (TOPMed) program was supported by the National Heart, Lung and Blood Institute (NHLBI). Metabolomics for "NHLBI TOPMed: “Multi-Ethnic Study of Atherosclerosis (MESA)” (phs001416) was performed at Broad Institute and Beth Israel Metabolomics Platform (HHSN268201600034I). We gratefully acknowledge the studies and participants who provided biological samples and data for TOPMed. Phase 1 TOPMed metabQTL results were generated in a collaboration between the TOPMed Metabolomics Working Group, the TOPMed Informatics Research Center, and the TOPMed parent studies contributing metabolite data and distributed to TOPMed investigators.

## Supplementary Materials

### MESA Sleep Ancillary Study

The Multi-Ethnic Study of Atherosclerosis (MESA) is a long-term study examining risk factors for heart disease in four ethnic groups. The full study design has been previously published(100). The study began in 2000 and recruited 6,814 adults with no clinical cardiovascular disease aged 45-84 years old from 6 field centres across the United States (Baltimore, MD; Chicago, IL; Los Angeles, CA; New York, NY; Saint Paul, MN; and Winston-Salem, NC). The fifth follow up (Exam 5) took place between April 2010 and February 2013 and included 4,077 participants. Among this group, 2,261 participants were included for the MESA Sleep Ancillary Study and asked to complete sleep questionnaires in addition to Polysomnography and actigraphy data collection. Among this group, 2,240 participants completed sleep questionnaires including ESS (11).

### Methods

Metabolomic analysis in the MESA study was performed for the blood samples of 2,640 participants from exam 5, of which 1,849 were part of the MESA Sleep Ancillary Study and completed the ESS questionnaire. Details regarding metabolomic quantification using the Broad Institute and Beth Israel Metabolomics Platform has been described elsewhere(26,101). Briefly, metabolite profiling utilized liquid chromatography tandem mass spectrometry (LC–MS) with positive ion mode for water-soluble metabolites and lipids. Raw data was processed using TraceFinder 3.1 and Progenesis QI. For negative ionization mode, an Agilent 1290 LC system coupled with a Waters XBridge Amide column and Agilent 6490 triple quadrupole mass spectrometer was employed. Isotope-labelled internal standards ensured MS sensitivity. Pooled plasma samples were interspersed for quality control. Metabolite identities were confirmed using authentic reference standards (26). In total, 4,380 metabolites were quantified from 1,868 exam 5 samples. Institutional Review Board approval was obtained at each study site and written informed consent was obtained from all participants.

### Mendelian Randomization metabQTL data

The Phase 1 Trans-Omics in Precision Medicine (TOPMed) metabQTL included metabolomics data from the Childhood Asthma Management Program (CAMP) (N = 787 using the Broad platform), The Genetic Epidemiology of Asthma in Costa Rica (CRA) study (N=1,758 using the Broad platform), Framingham Heart Study (FHS) (N = 3,021 using the Broad platform), MESA (n=998), Women’s Health Initiative (WHI) (N = 1,024 using the Broad platform and N= 545 using the Metabolon platform), Genetic Epidemiology of Chronic Obstructive Pulmonary Disease (COPDGene) Study (N=6,302 using the Metabolon platform), and the Subpopulations and Intermediate Outcome Measures in COPD Study (SPIROMICS) (N =1,924 using the platform) metabQTL summary statistics included variants with MAF ≥0.5%.

### Sample Preparation

Samples were prepared using the automated MicroLab STAR® system from Hamilton Company. Several recovery standards were added prior to the first step in the extraction process for QC purposes. To remove protein, dissociate small molecules bound to protein or trapped in the precipitated protein matrix, and to recover chemically diverse metabolites, proteins were precipitated with methanol under vigorous shaking for 2 min (Glen Mills GenoGrinder 2000) followed by centrifugation. The resulting extract was divided into five fractions: two for analysis by two separate reverse phase (RP)/UPLC-MS/MS methods with positive ion mode electrospray ionization (ESI), one for analysis by RP/UPLC-MS/MS with negative ion mode ESI, one for analysis by HILIC/UPLC-MS/MS with negative ion mode ESI, and one sample was reserved for backup. Samples were placed briefly on a TurboVap® (Zymark) to remove the organic solvent. The sample extracts were stored overnight under nitrogen before preparation for analysis.

### Ultrahigh Performance Liquid Chromatography-Tandem Mass Spectroscopy (UPLC-MS/MS)

All methods utilized a Waters ACQUITY ultra-performance liquid chromatography (UPLC) and a Thermo Scientific Q-Exactive high resolution/accurate mass spectrometer interfaced with a heated electrospray ionization (HESI-II) source and Orbitrap mass analyser operated at 35,000 mass resolution. The sample extract was dried then reconstituted in solvents compatible to each of the four methods. Each reconstitution solvent contained a series of standards at fixed concentrations to ensure injection and chromatographic consistency. One aliquot was analysed using acidic positive ion conditions, chromatographically optimized for more hydrophilic compounds. In this method, the extract was gradient eluted from a C18 column (Waters UPLC BEH C18-2.1×100 mm, 1.7 µm) using water and methanol, containing 0.05% perfluoropentanoic acid (PFPA) and 0.1% formic acid (FA). Another aliquot was also analysed using acidic positive ion conditions, however it was chromatographically optimized for more hydrophobic compounds. In this method, the extract was gradient eluted from the same afore mentioned C18 column using methanol, acetonitrile, water, 0.05% PFPA and 0.01% FA and was operated at an overall higher organic content. Another aliquot was analysed using basic negative ion optimized conditions using a separate dedicated C18 column. The basic extracts were gradient eluted from the column using methanol and water, however with 6.5mM Ammonium Bicarbonate at pH 8. The fourth aliquot was analysed via negative ionization following elution from a HILIC column (Waters UPLC BEH Amide 2.1×150 mm, 1.7 µm) using a gradient consisting of water and acetonitrile with 10mM Ammonium Formate, pH 10.8. The MS analysis alternated between MS and data-dependent MS^n^ scans using dynamic exclusion. The scan range varied slighted between methods but covered 70-1000 m/z. Raw data files are archived and extracted as described below.

### Data Extraction and Compound Identification

Raw data was extracted, peak-identified and QC processed using Metabolon’s hardware and software. These systems are built on a web-service platform utilizing Microsoft’s .NET technologies, which run on high-performance application servers and fibre-channel storage arrays in clusters to provide active failover and load-balancing. Compounds were identified by comparison to library entries of purified standards or recurrent unknown entities. Metabolon maintains a library based on authenticated standards that contains the retention time/index (RI), mass to charge ratio (*m/z)*, and chromatographic data (including MS/MS spectral data) on all molecules present in the library. Furthermore, biochemical identifications are based on three criteria: retention index within a narrow RI window of the proposed identification, accurate mass match to the library +/- 10 ppm, and the MS/MS forward and reverse scores between the experimental data and authentic standards. The MS/MS scores are based on a comparison of the ions present in the experimental spectrum to the ions present in the library spectrum. While there may be similarities between these molecules based on one of these factors, the use of all three data points can be utilized to distinguish and differentiate biochemicals. More than 3300 commercially available purified standard compounds have been acquired and registered into LIMS for analysis on all platforms for determination of their analytical characteristics. Additional mass spectral entries have been created for structurally unnamed biochemicals, which have been identified by virtue of their recurrent nature (both chromatographic and mass spectral). These compounds have the potential to be identified by future acquisition of a matching purified standard or by classical structural analysis.

### Metabolite Quantification and Data Normalization

Peaks were quantified using area-under-the-curve. For studies spanning multiple days, a data normalization step was performed to correct variation resulting from instrument inter-day tuning differences. Essentially, each compound was corrected in run-day blocks by registering the medians to equal one (1.00) and normalizing each data point proportionately.

## References

1. Gandhi KD, Mansukhani MP, Silber MH, Kolla BP. Excessive daytime sleepiness: A clinical review. Mayo Clin Proc. 2021 May;96(5):1288–301.

2. Pérez-Carbonell L, Mignot E, Leschziner G, Dauvilliers Y. Understanding and approaching excessive daytime sleepiness. Lancet. 2022 Sep 24;400(10357):1033–46.

3. Wang H, Lane JM, Jones SE, Dashti HS, Ollila HM, Wood AR, et al. Genome-wide association analysis of self-reported daytime sleepiness identifies 42 loci that suggest biological subtypes. Nat Commun. 2019 Aug 13;10(1):3503.

4. Bock J, Covassin N, Somers V. Excessive daytime sleepiness: an emerging marker of cardiovascular risk. Heart. 2022 Oct 28;108(22):1761–6.

5. Covassin N, Somers VK. Somnolence. Hypertension. 2016 Nov;68(5):1100–2.

6. Slater G, Steier J. Excessive daytime sleepiness in sleep disorders. J Thorac Dis. 2012 Dec;4(6):608–16.

7. Goldstein IB, Ancoli-Israel S, Shapiro D. Relationship between daytime sleepiness and blood pressure in healthy older adults. Am J Hypertens. 2004 Sep;17(9):787–92.

8. Connor J, Whitlock G, Norton R, Jackson R. The role of driver sleepiness in car crashes: a systematic review of epidemiological studies. Accident Analysis & Prevention. 2001 Jan;33(1):31–41.

9. Barfield R, Wang H, Liu Y, Brody JA, Swenson B, Li R, et al. Epigenome-wide association analysis of daytime sleepiness in the Multi-Ethnic Study of Atherosclerosis reveals African-American-specific associations. Sleep. 2019 Aug 1;42(8).

10. Hayes AL, Spilsbury JC, Patel SR. The Epworth score in African American populations. J Clin Sleep Med. 2009 Aug 15;5(4):344–8.

11. Chen X, Wang R, Zee P, Lutsey PL, Javaheri S, Alcántara C, et al. Racial/Ethnic Differences in Sleep Disturbances: The Multi-Ethnic Study of Atherosclerosis (MESA). Sleep. 2015 Jun 1;38(6):877–88.

12. Diallo I, Pak VM. Metabolomics, sleepiness, and sleep duration in sleep apnea. Sleep Breath. 2020 Dec;24(4):1327–32.

13. Sorlie PD, Avilés-Santa LM, Wassertheil-Smoller S, Kaplan RC, Daviglus ML, Giachello AL, et al. Design and implementation of the Hispanic Community Health Study/Study of Latinos. Ann Epidemiol. 2010 Aug;20(8):629–41.

14. Lavange LM, Kalsbeek WD, Sorlie PD, Avilés-Santa LM, Kaplan RC, Barnhart J, et al. Sample design and cohort selection in the Hispanic Community Health Study/Study of Latinos. Ann Epidemiol. 2010 Aug;20(8):642–9.

15. Siega-Riz AM, Sotres-Alvarez D, Ayala GX, Ginsberg M, Himes JH, Liu K, et al. Food-group and nutrient-density intakes by Hispanic and Latino backgrounds in the Hispanic Community Health Study/Study of Latinos. Am J Clin Nutr. 2014 Jun;99(6):1487–98.

16. Chiuve SE, Fung TT, Rimm EB, Hu FB, McCullough ML, Wang M, et al. Alternative dietary indices both strongly predict risk of chronic disease. J Nutr. 2012 Jun;142(6):1009–18.

17. Hispanic Community Health Study / Study of Latinos. HCHS/SOL Question by Question Instructions Physical Activity Form (PAE/PAS), Version A. 2007 May 12 [cited 2024 Jul 1]; Available from: https://sites.cscc.unc.edu/hchs/system/files/forms/UNLICOMMPhysicalActivityQxQ03062008.pdf

18. Radloff LS. The CES-D scale: A self-report depression scale for research in the general population. Appl Psychol Meas. 1977 Jun 1;1(3):385–401.

19. Ferreira R, Murray J. Spielberger’s State-Trait Anxiety Inventory: Measuring anxiety with and without an audience during performance on a stabilometer. Percept Mot Skills. 1983 Aug;57(1):15–8.

20. American Diabetes Association. Diagnosis and classification of diabetes mellitus. Diabetes Care. 2010 Jan;33 Suppl 1(Suppl 1):S62–9.

21. Levine DW, Kripke DF, Kaplan RM, Lewis MA, Naughton MJ, Bowen DJ, et al. Reliability and validity of the Women’s Health Initiative Insomnia Rating Scale. Psychol Assess. 2003 Jun;15(2):137–48.

22. Johns MW. A new method for measuring daytime sleepiness: the Epworth sleepiness scale. Sleep. 1991 Dec;14(6):540–5.

23. Redline S, Sotres-Alvarez D, Loredo J, Hall M, Patel SR, Ramos A, et al. Sleep-disordered breathing in Hispanic/Latino individuals of diverse backgrounds. The Hispanic Community Health Study/Study of Latinos. Am J Respir Crit Care Med. 2014 Feb 1;189(3):335–44.

24. Savin KL, Patel SR, Clark TL, Bravin JI, Roesch SC, Sotres-Alvarez D, et al. Relationships of Sleep Duration, Midpoint, and Variability with Physical Activity in the HCHS/SOL Sueño Ancillary Study. Behav Sleep Med. 2021;19(5):577–88.

25. Evans AM, DeHaven CD, Barrett T, Mitchell M, Milgram E. Integrated, nontargeted ultrahigh performance liquid chromatography/electrospray ionization tandem mass spectrometry platform for the identification and relative quantification of the small-molecule complement of biological systems. Anal Chem. 2009 Aug 15;81(16):6656–67.

26. Zhang Y, Ngo D, Yu B, Shah NA, Chen H, Ramos AR, et al. Development and validation of a metabolite index for obstructive sleep apnea across race/ethnicities. Sci Rep. 2022 Dec 16;12(1):21805.

27. Faquih T, van Smeden M, Luo J, le Cessie S, Kastenmüller G, Krumsiek J, et al. A workflow for missing values imputation of untargeted metabolomics data. Metabolites. 2020 Nov 26;10(12).

28. Zhang Y, Elgart M, Granot-Hershkovitz E, Wang H, Tarraf W, Ramos AR, et al. Genetic associations between sleep traits and cognitive ageing outcomes in the Hispanic Community Health Study/Study of Latinos. EBioMedicine. 2023 Jan;87:104393.

29. Li J, Ji L. Adjusting multiple testing in multilocus analyses using the eigenvalues of a correlation matrix. Heredity. 2005 Sep;95(3):221–7.

30. Friedman J, Hastie T, Tibshirani R. Sparse inverse covariance estimation with the graphical lasso. Biostatistics. 2008 Jul;9(3):432–41.

31. Epskamp S, Cramer AOJ, Waldorp LJ, Schmittmann VD, Borsboom D. qgraph: network visualizations of relationships in psychometric data. J Stat Softw. 2012;48(4).

32. Shannon P, Markiel A, Ozier O, Baliga NS, Wang JT, Ramage D, et al. Cytoscape: a software environment for integrated models of biomolecular interaction networks. Genome Res. 2003 Nov;13(11):2498–504.

33. Xia J, Psychogios N, Young N, Wishart DS. MetaboAnalyst: a web server for metabolomic data analysis and interpretation. Nucleic Acids Res. 2009 Jul;37(Web Server issue):W652–60.

34. Lu Y, Pang Z, Xia J. Comprehensive investigation of pathway enrichment methods for functional interpretation of LC-MS global metabolomics data. Brief Bioinformatics. 2023 Jan 19;24(1).

35. Goeman JJ, van de Geer SA, de Kort F, van Houwelingen HC. A global test for groups of genes: testing association with a clinical outcome. Bioinformatics. 2004 Jan 1;20(1):93–9.

36. Karnovsky A, Weymouth T, Hull T, Tarcea VG, Scardoni G, Laudanna C, et al. Metscape 2 bioinformatics tool for the analysis and visualization of metabolomics and gene expression data. Bioinformatics. 2012 Feb 1;28(3):373–80.

37. Ma H, Sorokin A, Mazein A, Selkov A, Selkov E, Demin O, et al. The Edinburgh human metabolic network reconstruction and its functional analysis. Mol Syst Biol. 2007 Sep 18;3:135.

38. Hemani G, Zheng J, Elsworth B, Wade KH, Haberland V, Baird D, et al. The MR-Base platform supports systematic causal inference across the human phenome. eLife. 2018 May 30;7.

39. Hemani G, Tilling K, Davey Smith G. Orienting the causal relationship between imprecisely measured traits using GWAS summary data. PLoS Genet. 2017 Nov 17;13(11):e1007081.

40. Rege J, Nanba AT, Auchus RJ, Ren J, Peng H-M, Rainey WE, et al. Adrenocorticotropin acutely regulates pregnenolone sulfate production by the human adrenal in vivo and in vitro. J Clin Endocrinol Metab. 2018 Jan 1;103(1):320–7.

41. Andersen ML, Bittencourt LRA, Antunes IB, Tufik S. Effects of progesterone on sleep: a possible pharmacological treatment for sleep-breathing disorders? Curr Med Chem. 2006;13(29):3575–82.

42. Kolatorova L, Vitku J, Suchopar J, Hill M, Parizek A. Progesterone: A Steroid with Wide Range of Effects in Physiology as Well as Human Medicine. Int J Mol Sci. 2022 Jul 20;23(14).

43. Abdrabou W, Dieng MM, Diawara A, Sermé SS, Almojil D, Sombié S, et al. Metabolome modulation of the host adaptive immunity in human malaria. Nat Metab. 2021 Jul;3(7):1001–16.

44. Li Y, Vgontzas AN, Fernandez-Mendoza J, Kritikou I, Basta M, Pejovic S, et al. Objective, but not subjective, sleepiness is associated with inflammation in sleep apnea. Sleep. 2017 Feb 1;40(2).

45. Rahman SA, Grant LK, Gooley JJ, Rajaratnam SMW, Czeisler CA, Lockley SW. Endogenous circadian regulation of female reproductive hormones. J Clin Endocrinol Metab. 2019 Dec 1;104(12):6049–59.

46. Liu PY, Reddy RT. Sleep, testosterone and cortisol balance, and ageing men. Rev Endocr Metab Disord. 2022 Dec;23(6):1323–39.

47. Kische H, Ewert R, Fietze I, Gross S, Wallaschofski H, Völzke H, et al. Sex Hormones and Sleep in Men and Women From the General Population: A Cross-Sectional Observational Study. J Clin Endocrinol Metab. 2016 Nov;101(11):3968–77.

48. Kelly MR, Yuen F, Satterfield BC, Auchus RJ, Gaddameedhi S, Van Dongen HPA, et al. Endogenous Diurnal Patterns of Adrenal and Gonadal Hormones During a 24-Hour Constant Routine After Simulated Shift Work. J Endocr Soc. 2022 Oct 26;6(12):bvac153.

49. Baker A, Simpson S, Dawson D. Sleep disruption and mood changes associated with menopause. J Psychosom Res. 1997 Oct;43(4):359–69.

50. Santoro N, Brown JR, Adel T, Skurnick JH. Characterization of reproductive hormonal dynamics in the perimenopause. J Clin Endocrinol Metab. 1996 Apr;81(4):1495–501.

51. Beverly Hery CM, Hale L, Naughton MJ. Contributions of the Women’s Health Initiative to understanding associations between sleep duration, insomnia symptoms, and sleep-disordered breathing across a range of health outcomes in postmenopausal women. Sleep Health. 2020 Feb;6(1):48–59.

52. Shahar E, Redline S, Young T, Boland LL, Baldwin CM, Nieto FJ, et al. Hormone replacement therapy and sleep-disordered breathing. Am J Respir Crit Care Med. 2003 May 1;167(9):1186–92.

53. Schüssler P, Kluge M, Yassouridis A, Dresler M, Held K, Zihl J, et al. Progesterone reduces wakefulness in sleep EEG and has no effect on cognition in healthy postmenopausal women. Psychoneuroendocrinology. 2008 Sep;33(8):1124–31.

54. Sigurðardóttir ES, Gislason T, Benediktsdottir B, Hustad S, Dadvand P, Demoly P, et al. Female sex hormones and symptoms of obstructive sleep apnea in European women of a population-based cohort. PLoS ONE. 2022 Jun 22;17(6):e0269569.

55. Driver HS, McLean H, Kumar DV, Farr N, Day AG, Fitzpatrick MF. The influence of the menstrual cycle on upper airway resistance and breathing during sleep. Sleep. 2005 Apr;28(4):449–56.

56. Lindberg E, Janson C, Gislason T, Björnsson E, Hetta J, Boman G. Sleep disturbances in a young adult population: can gender differences be explained by differences in psychological status? Sleep. 1997 Jun;20(6):381–7.

57. Rockwood K, Davis HS, Merry HR, MacKnight C, McDowell I. Sleep disturbances and mortality: results from the Canadian Study of Health and Aging. J Am Geriatr Soc. 2001 May;49(5):639–41.

58. Theorell-Haglöw J, Lindberg E, Janson C. What are the important risk factors for daytime sleepiness and fatigue in women? Sleep. 2006 Jun;29(6):751–7.

59. Wang M, He Y, Eisenman LN, Fields C, Zeng C-M, Mathews J, et al. 3beta - hydroxypregnane steroids are pregnenolone sulfate-like GABA(A) receptor antagonists. J Neurosci. 2002 May 1;22(9):3366–75.

60. Steiger A, Trachsel L, Guldner J, Hemmeter U, Rothe B, Rupprecht R, et al. Neurosteroid pregnenolone induces sleep-EEG changes in man compatible with inverse agonistic GABAA-receptor modulation. Brain Res. 1993 Jul 2;615(2):267–74.

61. Lancel M, Faulhaber J, Holsboer F, Rupprecht R. Progesterone induces changes in sleep comparable to those of agonistic GABAA receptor modulators. Am J Physiol. 1996 Oct;271(4 Pt 1):E763–72.

62. Gottesmann C. GABA mechanisms and sleep. Neuroscience. 2002;111(2):231–9.

63. Nielsen S. Benzodiazepines. Curr Top Behav Neurosci. 2017;34:141–59.

64. Friess E, Tagaya H, Trachsel L, Holsboer F, Rupprecht R. Progesterone-induced changes in sleep in male subjects. Am J Physiol. 1997 May;272(5 Pt 1):E885–91.

65. Jacobs MN, Lewis DFV. Steroid hormone receptors and dietary ligands: a selected review. Proc Nutr Soc. 2002 Feb;61(1):105–22.

66. Papandreou C. Polyunsaturated fatty acids in relation to sleep quality and depression in obstructive sleep apnea hypopnea syndrome. Omega-3 Fatty Acids in Brain and Neurological Health. Elsevier; 2014. p. 337–47.

67. Papandreou C, Camacho-Barcia L, García-Gavilán J, Hansen TT, Hjorth MF, Halford JCG, et al. Circulating metabolites associated with objectively measured sleep duration and sleep variability in overweight/obese participants: a metabolomics approach within the SATIN study. Sleep. 2019 May 1;42(5).

68. Peuhkuri K, Sihvola N, Korpela R. Diet promotes sleep duration and quality. Nutr Res. 2012 May;32(5):309–19.

69. Montgomery P, Burton JR, Sewell RP, Spreckelsen TF, Richardson AJ. Fatty acids and sleep in UK children: subjective and pilot objective sleep results from the DOLAB study--a randomized controlled trial. J Sleep Res. 2014 Aug;23(4):364–88.

70. Hansen AL, Dahl L, Olson G, Thornton D, Graff IE, Frøyland L, et al. Fish consumption, sleep, daily functioning, and heart rate variability. J Clin Sleep Med. 2014 May 15;10(5):567–75.

71. Xiao Q, Derkach A, Moore SC, Zheng W, Shu X-O, Gu F, et al. Habitual Sleep and human plasma metabolomics. Metabolomics. 2017 May;13(5).

72. Dallmann R, Viola AU, Tarokh L, Cajochen C, Brown SA. The human circadian metabolome. Proc Natl Acad Sci USA. 2012 Feb 14;109(7):2625–9.

73. Mustonen A-M, Nieminen P. Dihomo-γ-Linolenic Acid (20:3n-6)-Metabolism, Derivatives, and Potential Significance in Chronic Inflammation. Int J Mol Sci. 2023 Jan 20;24(3).

74. Campanini MZ, Guallar-Castillón P, Rodríguez-Artalejo F, Lopez-Garcia E. Mediterranean diet and changes in sleep duration and indicators of sleep quality in older adults. Sleep. 2017 Mar 1;40(3).

75. Castro-Diehl C, Wood AC, Redline S, Reid M, Johnson DA, Maras JE, et al. Mediterranean diet pattern and sleep duration and insomnia symptoms in the Multi-Ethnic Study of Atherosclerosis. Sleep. 2018 Nov 1;41(11).

76. Godos J, Ferri R, Caraci F, Cosentino FII, Castellano S, Galvano F, et al. Adherence to the Mediterranean Diet is Associated with Better Sleep Quality in Italian Adults. Nutrients. 2019 Apr 28;11(5).

77. Ferranti R, Marventano S, Castellano S, Giogianni G, Nolfo F, Rametta S, et al. Sleep quality and duration is related with diet and obesity in young adolescent living in Sicily, Southern Italy. Sleep Sci. 2016 Apr 14;9(2):117–22.

78. Lucki NC, Sewer MB. The interplay between bioactive sphingolipids and steroid hormones. Steroids. 2010 Jun;75(6):390–9.

79. Lucki NC, Sewer MB. Multiple roles for sphingolipids in steroid hormone biosynthesis. Subcell Biochem. 2008;49:387–412.

80. Zhang Y, Yu B, Qi Q, Azarbarzin A, Chen H, Shah NA, et al. Metabolomic profiles of sleep-disordered breathing are associated with hypertension and diabetes mellitus development. Nat Commun. 2024 Feb 28;15(1):1845.

81. Hiroi T, Imaoka S, Funae Y. Dopamine formation from tyramine by CYP2D6. Biochem Biophys Res Commun. 1998 Aug 28;249(3):838–43.

82. Burns C, Kidron A. Biochemistry, Tyramine. StatPearls. Treasure Island (FL): StatPearls Publishing; 2024.

83. Scammell TE. Overview of sleep: the neurologic processes of the sleep-wake cycle. J Clin Psychiatry. 2015 May;76(5):e13.

84. Monti J, Jantos H. The roles of dopamine and serotonin, and of their receptors, in regulating sleep and waking. Serotonin–dopamine interaction: experimental evidence and therapeutic relevance. Elsevier; 2008. p. 625–46.

85. Jouvet M. Sleep and serotonin: an unfinished story. Neuropsychopharmacology. 1999 Aug;21(2 Suppl):24S–27S.

86. Menon JML, Nolten C, Achterberg EJM, Joosten RNJMA, Dematteis M, Feenstra MGP, et al. Brain Microdialysate Monoamines in Relation to Circadian Rhythms, Sleep, and Sleep Deprivation - a Systematic Review, Network Meta-analysis, and New Primary Data. J Circadian Rhythms. 2019 Jan 14;17:1.

87. Vernia F, Di Ruscio M, Ciccone A, Viscido A, Frieri G, Stefanelli G, et al. Sleep disorders related to nutrition and digestive diseases: a neglected clinical condition. Int J Med Sci. 2021 Jan 1;18(3):593–603.

88. Mantas I, Vallianatou T, Yang Y, Shariatgorji M, Kalomoiri M, Fridjonsdottir E, et al. TAAR1-Dependent and -Independent Actions of Tyramine in Interaction With Glutamate Underlie Central Effects of Monoamine Oxidase Inhibition. Biol Psychiatry. 2021 Jul 1;90(1):16–27.

89. Schwartz MD, Black SW, Fisher SP, Palmerston JB, Morairty SR, Hoener MC, et al. Trace Amine-Associated Receptor 1 Regulates Wakefulness and EEG Spectral Composition. Neuropsychopharmacology. 2017 May;42(6):1305–14.

90. Schwartz MD, Canales JJ, Zucchi R, Espinoza S, Sukhanov I, Gainetdinov RR. Trace amine-associated receptor 1: a multimodal therapeutic target for neuropsychiatric diseases. Expert Opin Ther Targets. 2018 Jun 5;22(6):513–26.

91. Surendran P, Stewart ID, Au Yeung VPW, Pietzner M, Raffler J, Wörheide MA, et al. Rare and common genetic determinants of metabolic individuality and their effects on human health. Nat Med. 2022 Nov 10;28(11):2321–32.

92. Yap CX, Sidorenko J, Wu Y, Kemper KE, Yang J, Wray NR, et al. Dissection of genetic variation and evidence for pleiotropy in male pattern baldness. Nat Commun. 2018 Dec 20;9(1):5407.

93. Pickrell JK, Berisa T, Liu JZ, Ségurel L, Tung JY, Hinds DA. Detection and interpretation of shared genetic influences on 42 human traits. Nat Genet. 2016 Jul;48(7):709–17.

94. Pinilla L, Benítez ID, Gracia-Lavedan E, Torres G, Mínguez O, Vaca R, et al. Metabolipidomic Analysis in Patients with Obstructive Sleep Apnea Discloses a Circulating Metabotype of Non-Dipping Blood Pressure. Antioxidants (Basel). 2023 Nov 27;12(12).

95. Cole LK, Vance JE, Vance DE. Phosphatidylcholine biosynthesis and lipoprotein metabolism. Biochim Biophys Acta. 2012 May;1821(5):754–61.

96. Laird DA, Drexel H. Experimenting with Foods and Sleep I. Effects of Varying Types of Food in Offsetting Sleep Disturb-Ances Caused by Hunger Pangs and Gastric Distress— Children and Adults. J Am Diet Assoc. 1934 Jul;10(2):89–99.

97. Southwell PR, Evans CR, Hunt JN. Effect of a hot milk drink on movements during sleep. Br Med J. 1972 May 20;2(5811):429–31.

98. Chua EC-P, Shui G, Cazenave-Gassiot A, Wenk MR, Gooley JJ. Changes in Plasma Lipids during Exposure to Total Sleep Deprivation. Sleep. 2015 Nov 1;38(11):1683–91.

99. Yu Z, Zhai G, Singmann P, He Y, Xu T, Prehn C, et al. Human serum metabolic profiles are age dependent. Aging Cell. 2012 Dec;11(6):960–7.

100. Bild DE, Bluemke DA, Burke GL, Detrano R, Diez Roux AV, Folsom AR, et al. Multi-Ethnic Study of Atherosclerosis: objectives and design. Am J Epidemiol. 2002 Nov 1;156(9):871–81.

101. Paynter NP, Balasubramanian R, Giulianini F, Wang DD, Tinker LF, Gopal S, et al. Metabolic predictors of incident coronary heart disease in women. Circulation. 2018 Feb 20;137(8):841–53.

